# Cuffless Photoplethysmography Device for Continuous Monitoring of Blood Pressure

**DOI:** 10.1101/2025.02.08.25321920

**Authors:** Elias Hellou, Tamer Jamal, Elchanan Zuroff, Parvin Kalhor, Maria Delgado-Lelievre, Lilach O. Lerman, Amir Lerman, Erez Kachel, Ziad Zoghby

**Author notes:** Senior Co-Authorship. **Address for Correspondence:** Ziad Zoghby M.D. M.B.A. 200 First St SW Rochester, MN 55905.

## Abstract

**Introduction:** Accurate and convenient monitoring of BP is challenging and relies on cuff-based devices or in the postoperative and intensive care settings, on invasive measurements. The aim of this study was to prospectively evaluate the accuracy of BP measurements obtained from a novel, commercially available cuffless, non-invasive photoplethysmography (PPG)-based chest patch monitor compared to the reference standard invasive arterial pressure (IAP) monitoring, in patients after cardiac surgery.

**Methods:** This single center prospective study enrolled adults who underwent cardiac surgery. The PPG-based data were compared to IAP as part of standard of care. Bland-Altman plots and Pearson’s correlations were used to assess the accuracy between the two techniques.

**Results:** Ninety-six patients consented for the study. Mean age was 63.2±12.2 years (range 24 to 84), and 32 (33%) were women. Average monitoring time was 25.6±17.2. In total, we evaluated 78659 readings for systolic BP (SBP), 78818 for diastolic BP (DBP), and 92544 for HR analysis. These yielded correlation coefficients of r=0.959, 0.973, 0.966, and 0.962 for SBP, DBP, mean arterial pressure (MAP), and heart rate (HR), respectively. The Bland-Altman analysis showed a bias±SD of 0.1±4.8 mmHg for SBP; 0.4±2.1 mmHg for DBP; 0.26±2.6 mmHg for MAP, and 0.15±3.6 beats per minutes for HR. 95% of SBP, and 99.9% of DBP measurements were within 10 mmHg of the reference measurement.

**Conclusion:** Cuffless device offers a high level of accuracy of BP and HR, supporting the use of this novel noninvasive tool for continuous BP monitoring. Further studies are needed to validate those findings.

## Introduction

Blood pressure (BP) is a key physiological biomarker and its measurement is one of the most commonly performed medical procedures in the healthcare setting. Both hypertension and hypotension are associated with high morbidity and mortality. Hypertension, for example, is the leading modifiable risk factor for cardiovascular events^1^. Each 10 mm Hg reduction in systolic blood pressure (SBP) significantly reduces the risk of major cardiovascular events, coronary heart disease, stroke, and heart failure, and leads to a significant reduction in all-cause mortality^2^. Given the marked value of this parameter, over several decades, various techniques and devices have been developed for measuring BP^3^. The 2023 European Society of Hypertension (ESH)^4^ and 2017 American College of Cardiology/American Heart Association (ACC/AHA) hypertension^5^ guidelines both emphasize the importance of proper BP measurement. The BP measurement methods are traditionally divided into two main categories: direct measurement through invasive intra-arterial approaches for continuous monitoring of BP perioperatively or in the critical care setting; and non-invasive indirect measurement by creating arterial occlusion using cuff-based devices mostly used in the ambulatory setting. This later technique is based on the concept of Korotkoff sounds and measures the pressure using a sphygmomanometer, and more commonly now, using an automated oscillometric device that estimates BP based on a proprietary algorithm developed by the manufacturer. Recently, novel cuffless devices have been developed to noninvasively measure BP. Their clear advantage is their ability to provide frequent intermittent or continuous (an output period of more or under every 30 seconds, respectively) measurements in a non-invasive, convenient, and comfortable manner (with no need for arterial occlusion)^6,7^ which may facilitate BP monitoring in the inpatient and outpatient settings.

In the last few years, various Photoplethysmography (PPG) based cuffless devices have been approved for BP measurement (e.g. Biobeat^8^, LiveMetric^9^) in the United States or Europe (e.g. Aktiia). However, these devices are not endorsed yet by the scientific community and are not included in the American or European guidelines as acceptable alternatives for cuff-based devices. In 2022, the International Organization for Standardization (ISO) published its first validation standard (ISO 81060-3:2022) that is specific for continuous noninvasive sphyngomanometers^10^. In 2023, the European Society of Hypertension (ESH) published recommendations for the validation of intermittent cuffless BP measuring devices^11^.

Several commercially available cuffless-based devices have been evaluated in various clinical settings. For example, the Biobeat system (WP613, Biobeat Technology LTD, Petch Tikva, Israel) has been previously tested in the outpatient setting and found to provide comparable measurements to those obtained by a standard 24-hour ambulatory blood pressure monitor (ABPM)^12^. In a pilot study of BP monitoring after cardiac surgery, the device showed high level of accuracy compared to invasive intra-arterial pressure (IAP), however, this study included only 10 participants.^13^

The aim of this study was to establish the accuracy of a novel cuffless device (Biobeat) for continuous BP monitoring after cardiac surgery against a reference standard as an important step towards its clinical application. Specifically, the current study was designed to test the accuracy and the level of agreement between the BP measurements obtained by the cuffless PPG-based device and an IAP monitoring system, in patients in the Intensive Cardiac Care Unit (ICCU) after cardiac surgery. Because it is commercially available, this device may find widespread clinical use.

## Methods

This single center, prospective study recruited males and females 18-years or older admitted to the ICCU after cardiac surgery (Coronary Artery Bypass Grafting or valve surgery) at Chaim Sheba Medical Center, Ramat Gan, Israel, and had a concomitant intra-arterial (radial artery) catheter placed for post-operative BP monitoring per standard clinical care. Exclusion criteria were patients with severe postoperative shock state during the ICCU stay, pregnant women, patients with a lack of judgment/mental illness, and hospital personnel. The study protocol was approved by the institutional review board and registered in ClinicalTrials.gov ID NCT04071015.

Patients gave written informed consent preoperatively, and demographics and other relevant characteristics were collected. In addition, patients’ skin tone based on Fitzpatrick scale^14^ was recorded per FDA guidelines^15^ because skin tone may affect the accuracy of PPG-based devices. The study did not interfere with the operative flow. Postoperatively, patients were monitored in the ICCU; Biobeat (WP613 device from Biobeat Technology LTD, Petch Tikva, Israel) was attached to the chest wall following the manufacturer’s instructions. Continuous real-time readings of BP and heart rate (HR) were recorded simultaneously by both Biobeat and a standard IAP monitoring system (IntelliVue MX500 Patient Monitor, Philips Medical Systems). The monitoring was continued as long as IAP was clinically indicated, and up to five days (144 hours). Mean arterial pressure (MAP) was calculated from SBP and diastolic BP (DBP). Biobeat can produce one lead ECG tracing, but HR for analysis was intentionally obtained from the cuffless device as derived from the cuffless PPG-based system. All data were processed and analyzed post hoc. No information from the Biobeat was available to medical personnel at the time of the study, or used in clinical decision-making. Data from Biobeat was available every 5 seconds, data recorded by IAP was available every 1 minute, and simultaneous measurements were used for comparison. ICCU monitors were connected to IAP as a standard measure in ICCU patients, and calibrated according to instructions of use and according to the clinical practice^16^. IAP was calibrated periodically per protocol by bedside registered nurses, at the beginning of the monitoring and at least once every 8 hours. Valid ranges for HR were considered 35 to 155 beats per minute (bpm), for SBP 60 to 300 mmHg, and for DBP 30 to 150 mmHg due to the IAP calibration ranges and limitations.

### Data Analysis

Bland-Altman plots and Pearson’s correlations were used to assess the accuracy and degree of agreement between measurement techniques for each index (SBP, DBP, MAP, and HR). Significant differences were compared using a mixed-effects repeated measures ANOVA approach and deemed statistically significant where *p* < 0.05 with differences described as mean±standard deviation. All measurements for each subject were averaged and compared for the entire dataset. A heatmap was plotted to show the number of measurements within 4 groups of precision (0-5, 6-10, 11-15, and above 15 mmHg for BP and beats-per-minute for HR.

## Results

Ninety-six (96) patients were recruited in the study. **Figure 1** shows the study flow. Among them, 4 patients had no BP log file for IAP due to technical issues, and 4 patients had severe shock state or returned to the operating room with removal of the patch. These patients were excluded due to out-of-range IAP readings (low BP) or ventricular tachycardia or fibrillation, affecting calibration limits. For the BP, 88 patients were included in the final analysis, among who 80 were treated with inotropic agents during the monitoring period. **Table 1** shows the demographics and baseline characteristics of the patients. The average age was 63.2±12.2 years (range: 24-84), and 32 (33%) were women. Average monitoring time was 25.6±17.2 hours (range: 8-142). In total, 78659 readings were obtained for SBP, 78818 readings for DBP, and 78212 for MAP. For the HR analysis, data from 92 patients were included with a total of 92544 readings. Of these, 4695 readings were obtained from two permanent pacemaker-dependent patients.

**Figure 1:**
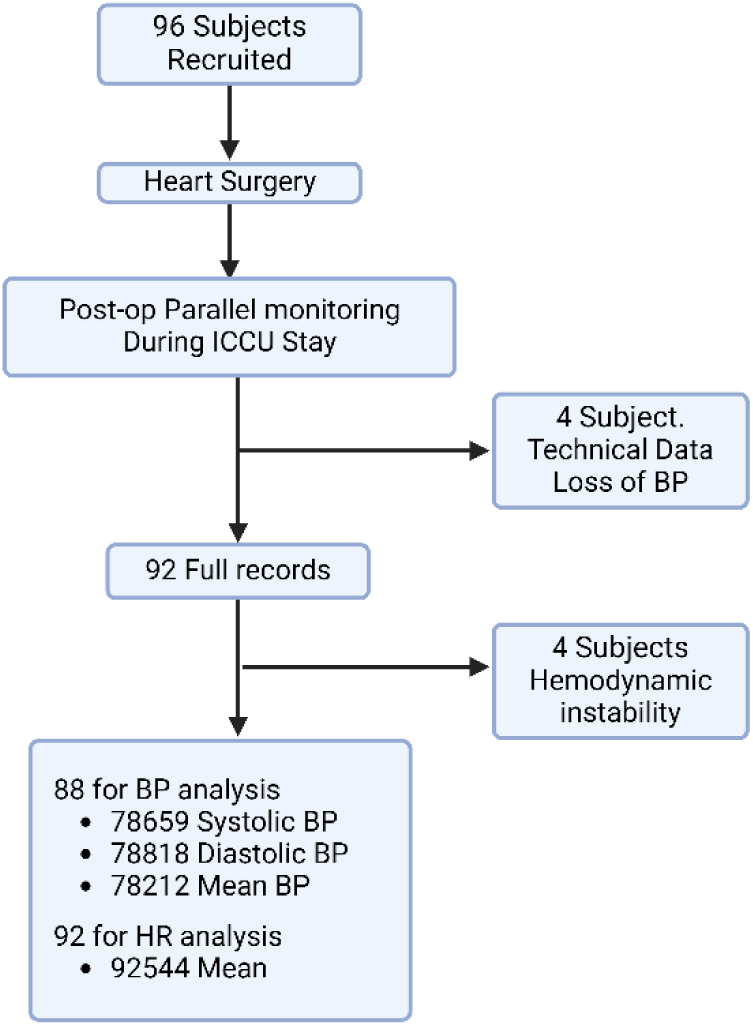
Experimental design. Blood pressure (BP); Intensive Cardiac Care Unit (ICCU); and Heart rate (HR)

**Table 1:** Demographics and patient characteristics and blood tests at baseline.

| **Patient Characteristics** | (N=96) |
| --- | --- |
| Age [years; Mean±SD] | 63.2±12.2 |
| Range [Min, Max] | [24.0, 84.0] |
| Females | 32 (33%) |
| Height [m; Mean±SD] | 1.69±0.09 |
| Weight [kg; Mean±SD] | 78.3±15.4 |
| BMI [kg/m<sup>2</sup>; Mean±SD] | 27.4±4.89 |
| Current Smokers | 19 (19.8%) |
| Obesity | 34 (35.4%) |
| Diabetes mellitus | 31 (32.3%) |
| Congestive heart disease | 11 (11.5%) |
| Hypertension | 60 (62.5%) |
| Ischemic heart disease | 40 (41.7%) |
| Valvular disease | 49 (51.0%) |
| Monitoring duration [hours; Mean±SD] | 25.6±17.2 |
| Median hours [Min, Max] | 22.0 [8.00, 142] |
| **Blood tests at baseline [Mean±SD]** |  |
| Lactate [mg/dL] | 16.7±10.0 |
| Base Excess [mEq/L] | -3.05±3.02 |
| HCO<sub>3</sub> [mEq/L] | 22.1±2.7 |
| pO<sub>2</sub> [mmHg] | 151±64 |
| pCO<sub>2</sub> [mmHg] | 40.3±7.9 |
| pH | 7.36±0.06 |
| Glucose [mg/dL] | 150±38.5 |
| Creatinine [mg/dL] | 1.22±1.32 |
| Platelets [1000/μL] | 180±57.5 |
| Hemoglobin [g/dL] | 11.5±1.7 |
| White blood cells [$10^3$/mm<sup>3</sup>] | 15.4±5.8 |

Standard deviation (SD) and Body mass index (BMI)

**Figure 2 and Table 2** show an excellent correlation between all the measurements obtained by the cuffless blood pressure (SBP, DBP, MAP, and HR) and the IAP monitoring system. The correlation coefficient was 0.959, 0.973, and 0.962 for SBP, DBP, and HR, respectively. The Bland-Altman analysis and plot show a bias±SD of 0.1±4.8 mmHg for SBP, 0.4±2.1 mmHg for DBP, and 0.15±3.6 bpm for HR. The excellent correlation was maintained across the BP continuum with no signs of measurement drift. Further analyses stratified by the different skin tones according to the Fitzpatrick scale (1 to 6) were consistent with the main results (**Table 3**).

**Figure 2:**
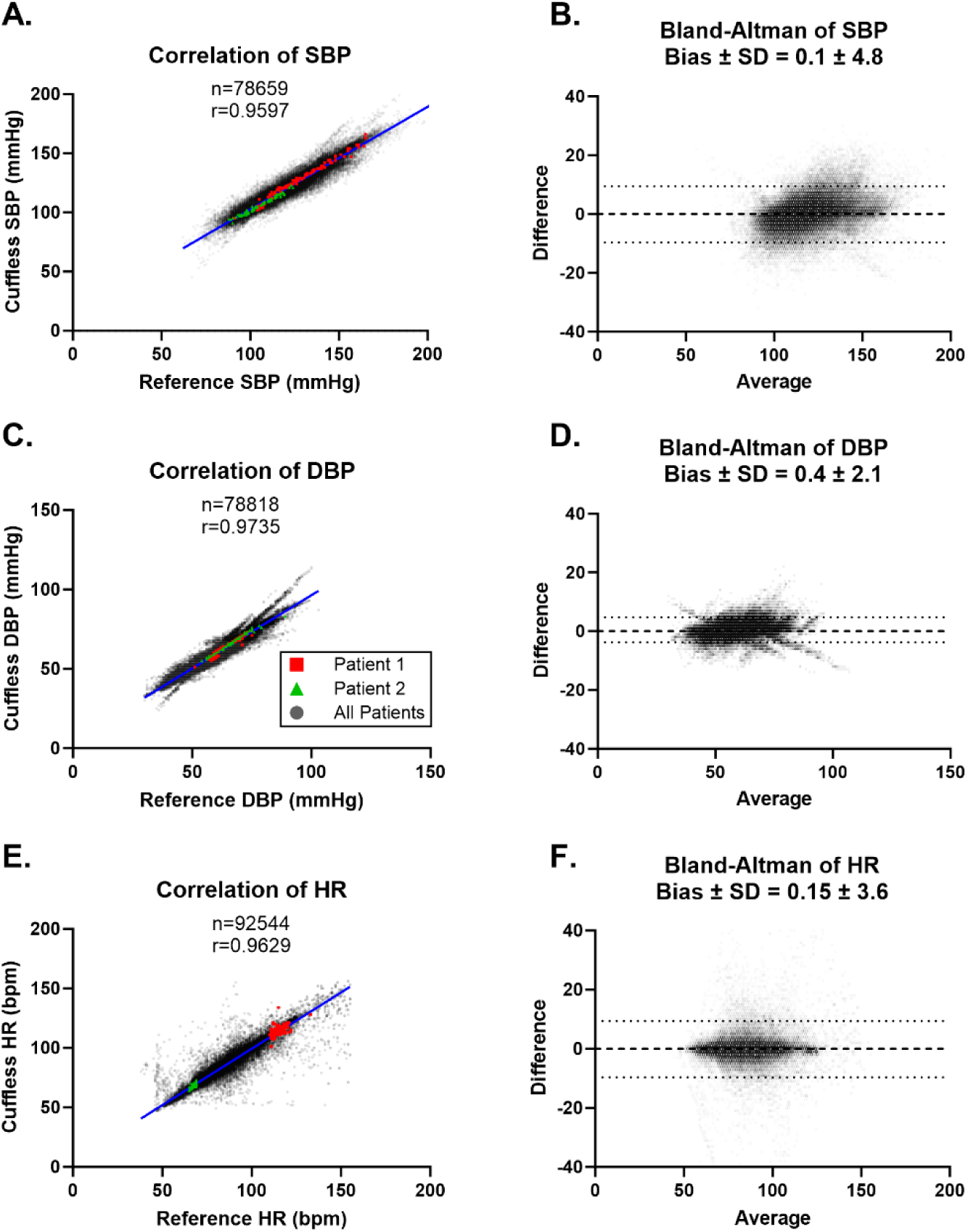
(A) The correlation of systolic BP between the tested device and IAP measurement, (B) a Bland-Altman scatterplot displaying the tested device and IAP systolic BP difference vs their average; (C) & (D) for diastolic BP; (E) & (F) for heart rate. The red and green datapoints are illustrative only and derived from two randomly selected patients. Systolic blood pressure (SBP); Diastolic blood pressure (DBP); Mean arterial pressure (MAP); Heart rate (HR); and standard deviation (SD)

**Table 2:** Bias and correlation of BP and HR measured by the Biobeat device and IAP.

|  | Bland-Altman<br>Bias±SD | Pearson r | Number of<br>paired<br>measurements | Difference<br>≤5 | Difference<br>≤10 | Difference<br>≤15 |
| --- | --- | --- | --- | --- | --- | --- |
| SBP, mmHg | 0.1±4.8 | 0.960 | 78659 | 80% | 95% | 99% |
| DBP, mmHg | 0.4±2.1 | 0.974 | 78818 | 97% | 99.9% | 99.9% |
| MAP, mmHg | 0.26±2.6 | 0.967 | 78212 | 94% | 99.6% | 99.9% |
| HR, bpm | 0.15±3.6 | 0.963 | 92544 | 95% | 98% | 99% |

Systolic blood pressure (SBP); Diastolic blood pressure (DBP); Mean arterial pressure (MAP); Heart rate (HR); Standard deviation (SD), and beats per minute (bpm)

**Table 3.** shows the correlation and bias of the systolic blood pressure of the Biobeat device according to subjects’ skin tone based on the Fitzpatrick scale (1-6).

| **Skin tone**<br>**(Fitzpatrick**<br>**scale)** | **Number (%)**<br>**N=96** | **Number**<br>**of pairs** | **Pearson r** | **P value** | **Bias±SD** | **95% Limits of**<br>**Agreement** |
| --- | --- | --- | --- | --- | --- | --- |
| **I** | 17 (17.7%) | 15718 | 0.970 | <0.0001 | 0.04±4.56 | -8.891 to 8.963 |
| **II** | 16 (16.7%) | 11282 | 0.950 | <0.0001 | -0.16±4.48 | -8.933 to 8.621 |
| **III** | 22 (22.9%) | 14861 | 0.958 | <0.0001 | -0.09±5.24 | -10.36 to 10.17 |
| **IV** | 15 (15.6%) | 12533 | 0.952 | <0.0001 | -0.07±4.79 | -9.454 to 9.317 |
| **V** | 14 (14.6%) | 13335 | 0.953 | <0.0001 | -0.01±4.92 | -9.656 to 9.634 |
| **VI** | 10 (10.4%) | 8007 | 0.958 | <0.0001 | -0.04±5.24 | -10.31 to 10.24 |
| **NA** | 2 (2.1%) |  |  |  |  |  |

SD, standard deviation; NA, not available

We conducted a further analysis and compared each participant’s average BP and HR obtained by the Biobeat and IAP systems. Figure 3 shows significant correlations (r=0.99 for SBP, DBP, and HR) and agreement (bias of 0.3±1.3 and 0.6±0.7mmHg for SBP and DBP, respectively). Figure 4 illustrates in a heat map the BP and HR bias based on four groups: bias of 0 to 5, 6 to 10, 11 to 15, and >15 mmHg or bpm for BP and HR, respectively. Lastly, Figure 5 shows representative examples illustrating that the changes in BP and HR measured by IAP were matched by Biobeat. In Figure 5b, a drift between IAP and Biobeat was due to a delay in the IAP calibration that resolved after calibration. Among the sub-group of patients (n=2) with permanent pacemakers, the correlation was excellent between cuffless BP and the invasive IAP readings with r=0.949 for SBP and 0.974 for DBP (Supplemental Figure S1).

**Figure 3:**
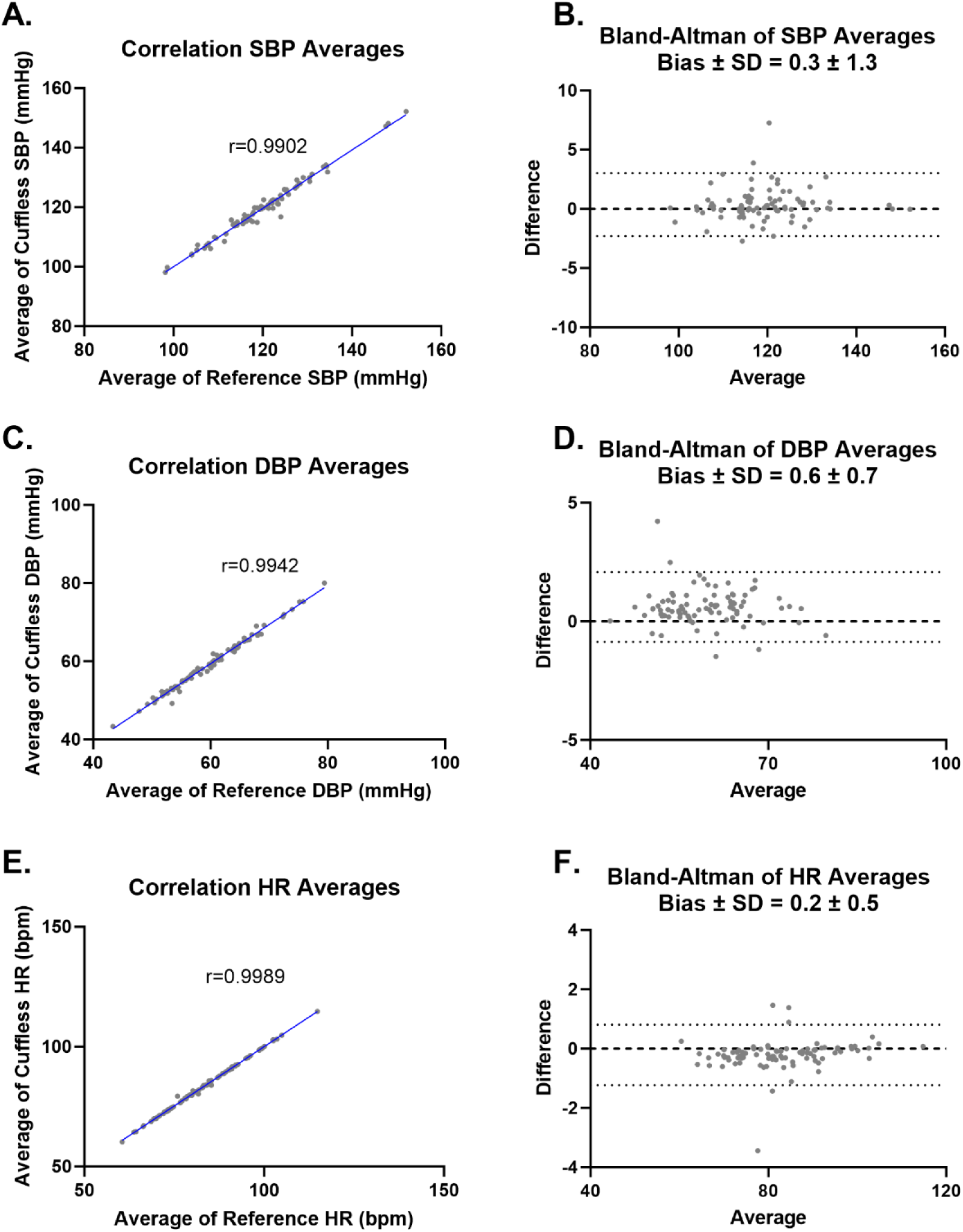
shows (A) classic presentation of the correlation of the average of systolic BP for each patient, as obtained from tested device and IAP, (B) Bland-Altman scatterplot displaying the tested device patient’s average and IAP average for systolic BP difference vs their average; (C) & (D) for diastolic BP, and (E) & (F) for heart rate.

**Figure 4:**
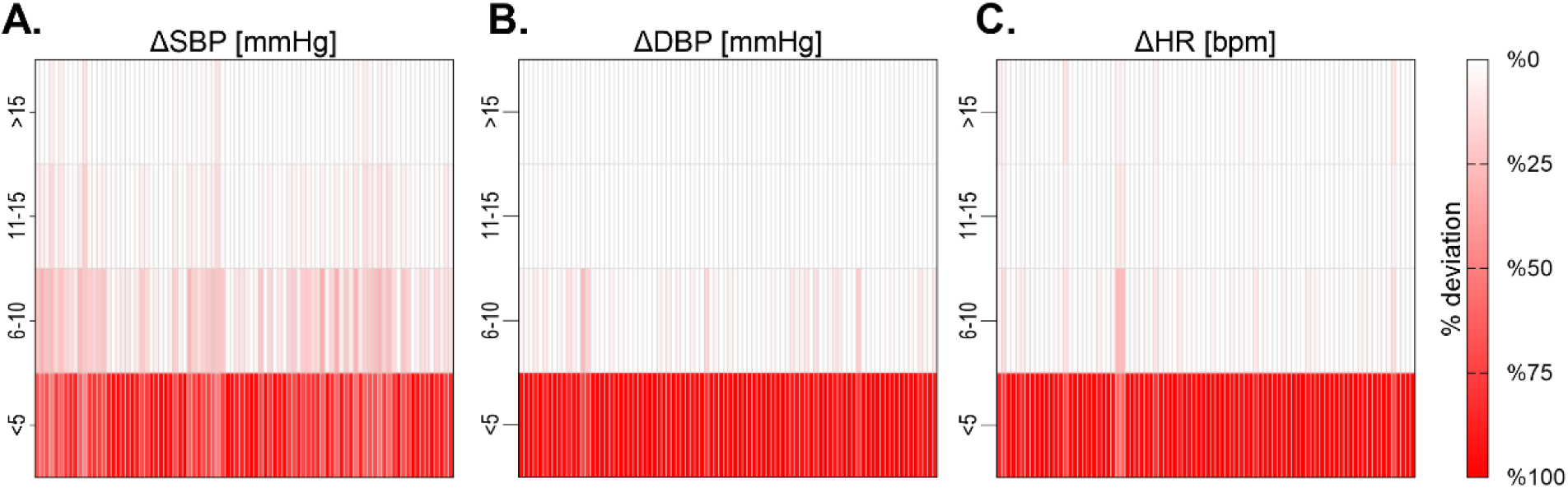
Distribution of bias within each participant. (A) represents systolic BP, (B) diastolic BP, and (C) heart rate. The differences were reported with-in 4 groups, with differences of 0 to 5, 6 to 10, 11 to 15, and more than 15 units (mmHg for BP and bpm for HR). The darker red color indicates a higher proportion of measurements that fall within that group. 86% of SBP, and 100% of DBP, measurements were within 10 mmHg of the reference measurement, and 95.4% of HR were within 10 bpm difference of reference measurement.

**Figure 5:**
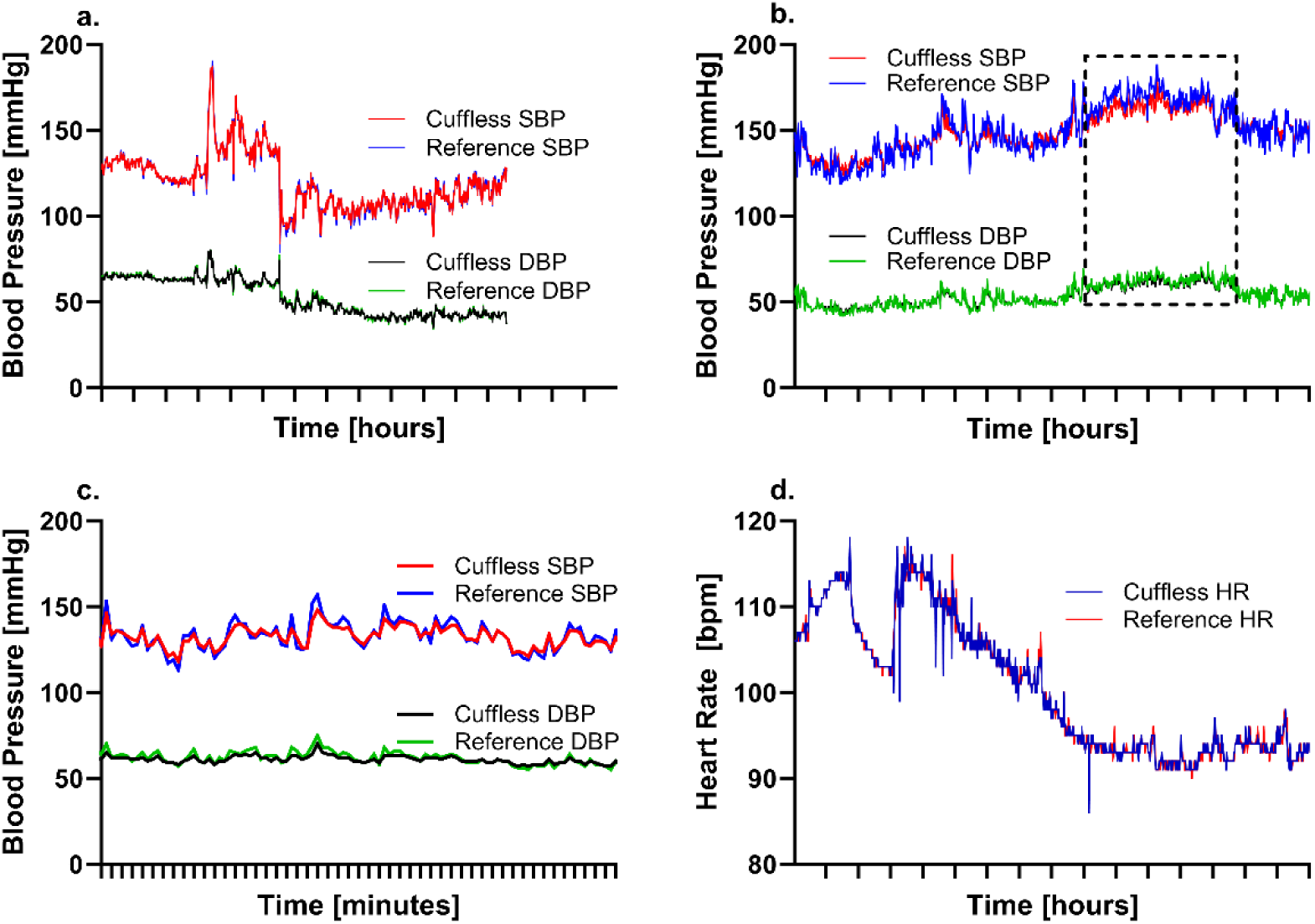
representative examples illustrating that the changes in BP and HR measured by IAP were matched by Biobeat over time. (A) shows SBP and DBP over time during thirteen hours of monitoring. (B) is similar to (A) for another patient, the box shows a drift between IAP and Biobeat due to a delay in the IAP calibration that resolved after calibration. (C) shows SBP and DBP over one hour. (D) shows HR over time during sixteen hours of monitoring for a randomly selected patient. Systolic blood pressure (SBP); Diastolic blood pressure (DBP); and Heart rate (HR)

## Discussion

The current study demonstrates excellent correlations, minimal bias and a high level of agreement in BP and HR measurements between the Biobeat cuffless PPG-based device and IAP. These results were consistent regardless of skin tone. These observations support the use of the Biobeat for noninvasive and convenient measurements of BP in patients in the ICCU.

The result of this study confirms the findings of a previous pilot study using the same device^13^ and adds to the accumulating evidence in support of cuffless BP devices^17–22^. Most published studies to date have evaluated cuffless devices for “intermittent” ambulatory use (compared with snapshot readings or 24-hours ABPM) rather than “continuous” monitoring after surgery or in the intensive care unit and included a small number of participants. This hinders the ability to validate the results in various populations with different skin tones. The current standards, however, used to validate invasive or cuff-based devices are not appropriate for cuffless devices due to the nature of this novel technology^6^. Because the pace of regulation of these devices did not keep up with the speed of innovation, prior studies have used traditional validation protocols^23^. More recently, consensus on standards by which cuffless BP devices can be tested for accuracy have been developed. Therefore, we expect in the future an acceleration in the adoption of cuffless devices assuming they pass the formal validation process^11,24^. Since cuffless BP devices estimate BP rather than directly or indirectly measure it, one of the main concerns has been the ability to track BP changes. Our study did demonstrate that BP and HR data from Biobeat matched very well those from the IAP as illustrated in randomly selected patients (Figure 5).

BP measurement is critical for clinical decision making both in the acute inpatient setting during hemodynamic monitoring and in the outpatient setting for long term management of hypertension. Yet, the accuracy and ease of obtaining BP measurements is problematic in clinical practice for various reasons (patient, healthcare system, and health care provider or care team factors) that are compounded by time constraints. This can contribute to misdiagnosis or diagnostic uncertainty potentially leading to inappropriate clinical decision making such as under or over treatment or therapeutic inertia. A major advantage of hemodynamic monitoring using PPG technology is its non-invasive nature, wireless set up, ease of operation with minimal training, and the lower risk of complications. While this study evaluated the performance of a cuffless device in the hospital, the technology has a wider application in the outpatient setting. Hypertension is most often diagnosed according to BP levels during office visits (intermittent snapshots) with a cuff-based device, or ideally by ABPM (intermittent recurrent pattern) with a timed cuff-based device. However, ABPM is not widely available and when available is not practical or convenient for ongoing monitoring. Thus, home BP measurements using a reliable technique and valid home BP device is now recommended by most major hypertension guidelines^4,5^. If proven to be accurate, reproducible over time, and able to track BP changes correctly (diurnal variation, antihypertensive drug effects, stress/activity), cuffless BP devices will transform the monitoring and management of hypertension, regardless of skin tone. The ability of this emerging technology to improve clinical outcomes, however, will need to be determined.

### Limitations

Our study has several limitations. First, this is a single-center study that assessed patients during recovery after cardiac surgery, and thus the results applicability to other populations and settings needs to be determined. Further studies to evaluate the ability of PPG-based devices to track BP should be performed in other settings. Second, this study was designed before the consensus guidelines for validating pulseless BP devices were published. These guidelines have more stringent recommendations, and future studies should be planned and conducted accordingly. As stated earlier, one of the main validation criteria in these guidelines is to demonstrate the ability of these devices to track BP changes over time. The results of this study are reassuring as the changes in BP matched well between Biobeat and IAP. Third, Biobeat can be used as a “continuous” or “intermittent” cuffless device, but each mode requires a different validation process depending on the intended use. In this study, the comparison was done with IAP which is the gold standard for continuous monitoring, however, this does not apply for intermittent ambulatory use in which the gold standard comparator is a 24-hour ABPM or manual auscultatory by trained and certified personnels, using a validated cuff device. Fourth, the cuffless BP patch can generate HR readings in two different ways: one that come from the single-lead ECG tracing, and the other one from PPG sensor that relies on BP peaks. The HR analysis was based on the PPG-based source, while the IAP monitor derived the HR reading from the ECG tracing in the patient monitoring system. Nevertheless, the HR correlation and agreement between the PPG device and IAP monitor was excellent. Last, even the gold-standard (IAP system) is not without flaws. IAP needs frequent calibrations, especially when changing the bed level, raising the head of the bed, and with posture changes, which can affect the reference BP readings and the interpretation of the results.

## Conclusion

The tested cuffless device (Biobeat) offers a high level of accuracy and agreement for BP and HR compared to data obtained by IAP monitoring in post-cardiac surgery patients. Further studies are needed to validate these findings in various populations and clinical settings according to the recently adopted validation protocols for pulseless devices.

Non-standard Abbreviations and Acronyms:

## Data Availability

All data produced in the present study are available upon reasonable request to the authors

## Ambulatory Blood Pressure Monitoring ABPM

BPM: Beats Per Minute
BP: Blood Pressure
BMI: Body Mass Index
CABG: Coronary Artery Bypass Grafting
DBP: Diastolic Blood Pressure
ECG: Electrocardiogram
FDA: United States Food and Drug Administration
HR: Heart Rate
IAP: Invasive Arterial Pressure
ICCU: Intensive Cardiac Care Unit
MAP: Mean Arterial Pressure
mmHg: Millimeters of Mercury
PPG: Photoplethysmography
SD: Standard Deviation
SBP: Systolic Blood Pressure

## Acknowledgments

Special thanks to all the staff, nurses and doctors who took part in performing this study; to the patients who enabled this study to be performed and consented to the study during the stressful event of heart surgery.

## Figures Legends

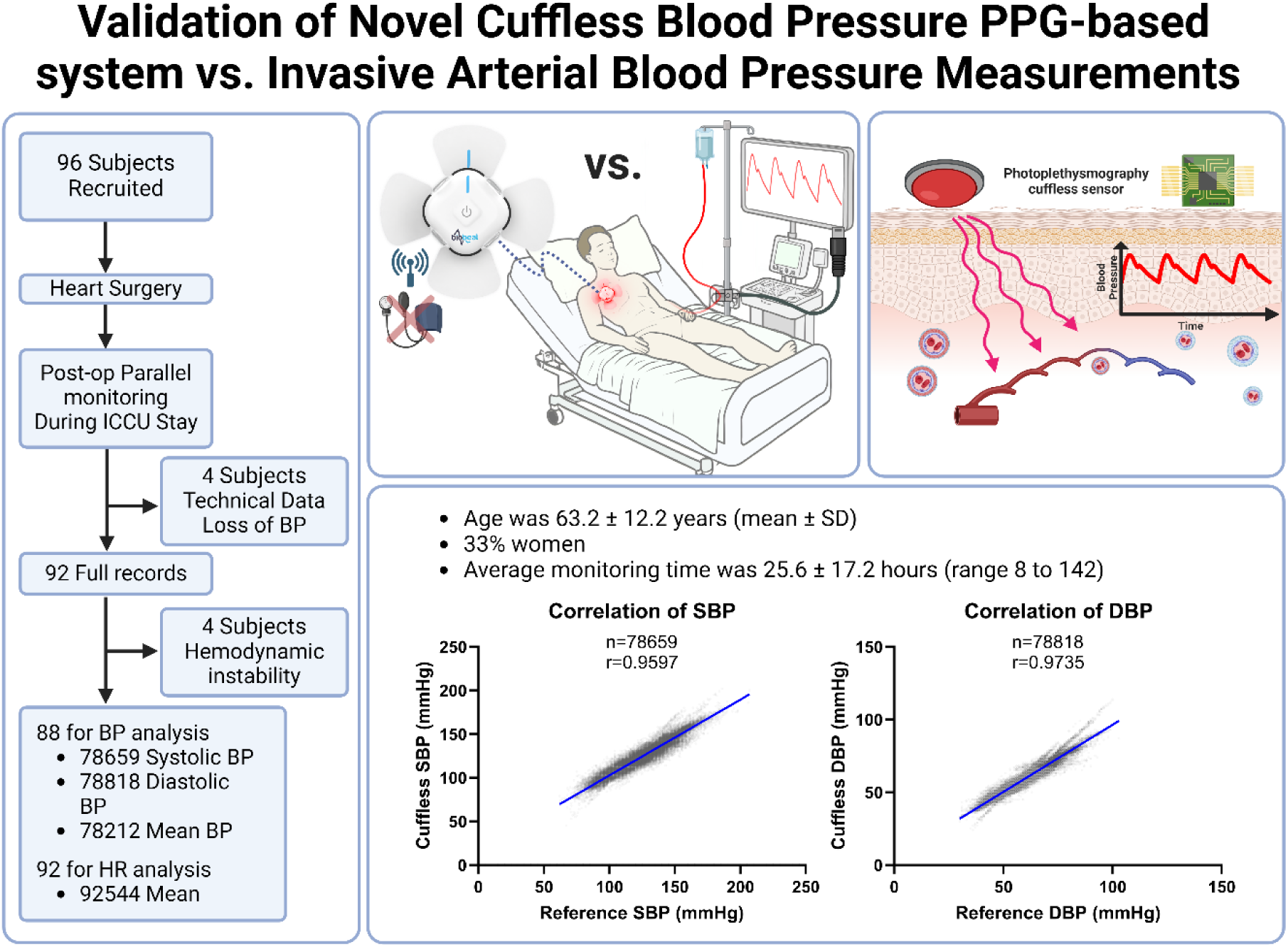
**Central Illustration**, demonstrating (A) patient’s flow chart; (B) Photoplethysmography sensor principals of action; (C) Study design, parallel measurement of IAP and cuffless device for patients in ICCU; and (D) the main results of the study. Blood pressure (BP); Systolic blood pressure (SBP); Diastolic blood pressure (DBP); Mean arterial pressure (MAP); and Heart rate (HR)

**Supplemental Figure S1:**
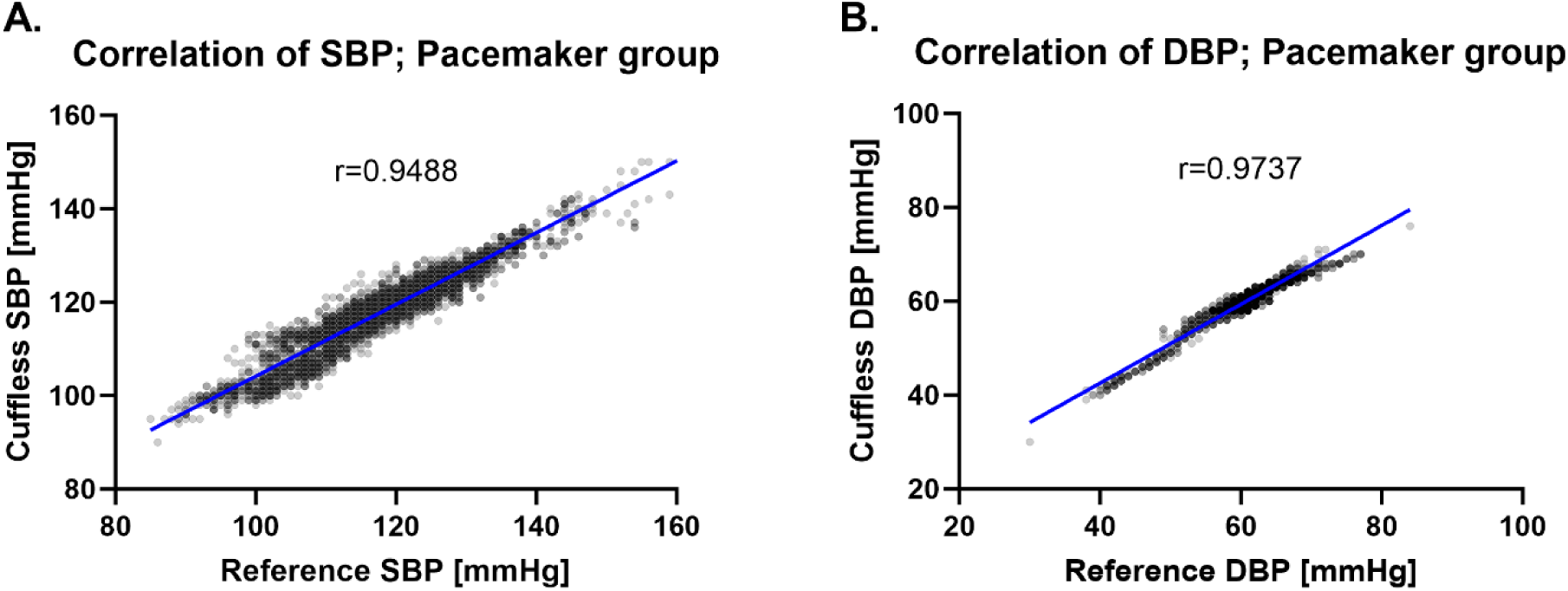
Correlation of BP measurements among the sub-group of patients with pacemakers also shows an excellent correlation between cuffless BP as compared to the invasive IAP readings.

